# Environmental profiles of urban living relate to regional brain volumes and symptom groups of mental illness through distinct genetic pathways

**DOI:** 10.1101/2022.09.08.22279549

**Authors:** Jiayuan Xu, Nana Liu, Elli Polemiti, Liliana Garcia Mondragon, Jie Tang, Xiaoxuan Liu, Tristram Lett, Le Yu, Markus Noethen, Chunshui Yu, Andre Marquand, Gunter Schumann

## Abstract

The majority of people worldwide live in cities, yet how urban living affects brain and mental illness is scarcely understood. Urban lives are exposed to a a wide array of environmental factors that may combine and interact to influence mental health. While individual factors of the urban environment have been investigated in isolation, to date no attempt has been made to model how the complex, real life exposure to living in the city relates to brain and mental illness, and how it is moderated by genetic factors. Using data of over 150,000 participants of the UK Biobank, we carried out sparse canonical correlation analyses (sCCA) to investigate the relation of urban living environment with symptoms of mental illness. We found three mental health symptom groups, consisting of affective, anxiety and emotional instability symptoms, respectively. These groups were correlated with distinct profiles of urban environments defined by risk factors related to social deprivation, air pollution and urban density, and protective factors involving green spaces and generous land use. The relations between environment and symptoms of mental illness were mediated by the volume of brain regions involved in reward processing, emotional processing and executive control, and moderated by genes regulating stress response, neurotransmission, neural development and differentiation, as well as epigenetic modifications. Together, these findings indicate distinct biological pathways by which different environmental profiles of urban living may influence mental illness. Our results also provide a quantitative measure of the contribution of each environmental factor to brain volume and symptom group. They will aid in targeting and prioritizing important decisions for planning and public health interventions.

## Introduction

More than 50% of the world population lives in an urban area, and it is estimated that by 2050, two-thirds of the world population will live in cities^1^. This dramatic increase in urbanization means that living environments are going through drastic transformations: Lives in urban areas are led in increasingly higher density residential and commercial buildings^1^, concomitant reduced access to green areas^2^, increased exposures to potentially harmful substances^3^ and more stressful social conditions^4^. At the same time urbans residents potentially benefit from better infrastructure and more work opportunities than rural dwellers^1^.

The impact of the urban living environment on mental health, is not well understood. Thus far there is evidence of urban-rural differences in the prevalence of psychiatric disorders^5^, and recent research has confirmed that individuals living in urban environments are at a higher risk of experiencing various mental health issues. The most prevalent of these issues are increased emotional problems, including symptoms of depression and anxiety^5-8^. In addition to the investigation of a general and complex environmental factor as urbanicity, there have been investigations of isolated environmental factors relevant to urban living, such as greenspaces ^9,10^, socio-economic status^11^ and others^12^. But these isolated factors have not been considered in the wider environmental context that characterises a living environment. In order to develop targeted prevention and intervention programmes ranging from urban planning to individual psychosocial coping programmes, it is neither sufficient to regard urbanicity as one risk factor, nor is it sufficient to focus on single isolated environmental factors alone. The urban environment, as any other living environment, is composed of simultaneous interacting factors, which may form profiles or signatures that together can reduce or increase risk for mental illness^13,14^.

The relation of symptoms of mental illness and brain structure with the exposure to environmental profiles of multiple factors composing a living environment are currently unknown, either in urban settings or otherwise. Furthermore, exposure to environmental adversity does not result in a uniform response, but individual differences have been well documented^15^. Genetic variations are known to be one important source of such individual differences^16^. In fact, activity of biological pathways, such as the stress response pathway or epigenetic modifications that are known mediators of the effect of stressful environmental stimuli on brain and mental illness have been shown to vary depending on particular genotypes^17^.

In this paper, we identify environmental profiles of urban living and relate them to groups of mental illness symptoms. We aim to understand what combinations of environmental profiles are most relevant for these emotional problems, and how within these combinations each single factor contributes to risk or resilience. We also identify regional brain areas that mediate the effect of different environmental profiles on mental illness symptoms groups. To discover moderators that may underlie individual differences in response to adverse environmental profiles, we investigate genetic variations derived from genome-wide analyses of symptoms of mental illness and test them for moderation of the regional brain volumes correlated to environmental adversity. To this end, we carried out four analysis steps: (1) a two-way sparse canonical correlation analysis (sCCA) to identify environmental profiles and their related symptom groups of mental illness. (2) a genome-wide association study (GWAS) of the symptom groups identified, followed by the computation of gene scores of GWAS-significant genes. (3) a three-way multiple-sparse canonical correlation analysis (msCCA) relating urban living environmental profiles and symptom groups with regional brain volume. (4) a moderated mediation analysis to characterize the mediation of environmental profiles and symptom groups by regional brain volume, and its moderation by gene scores (Figure 1 and Supplementary Figure 1).

**Figure 1:**
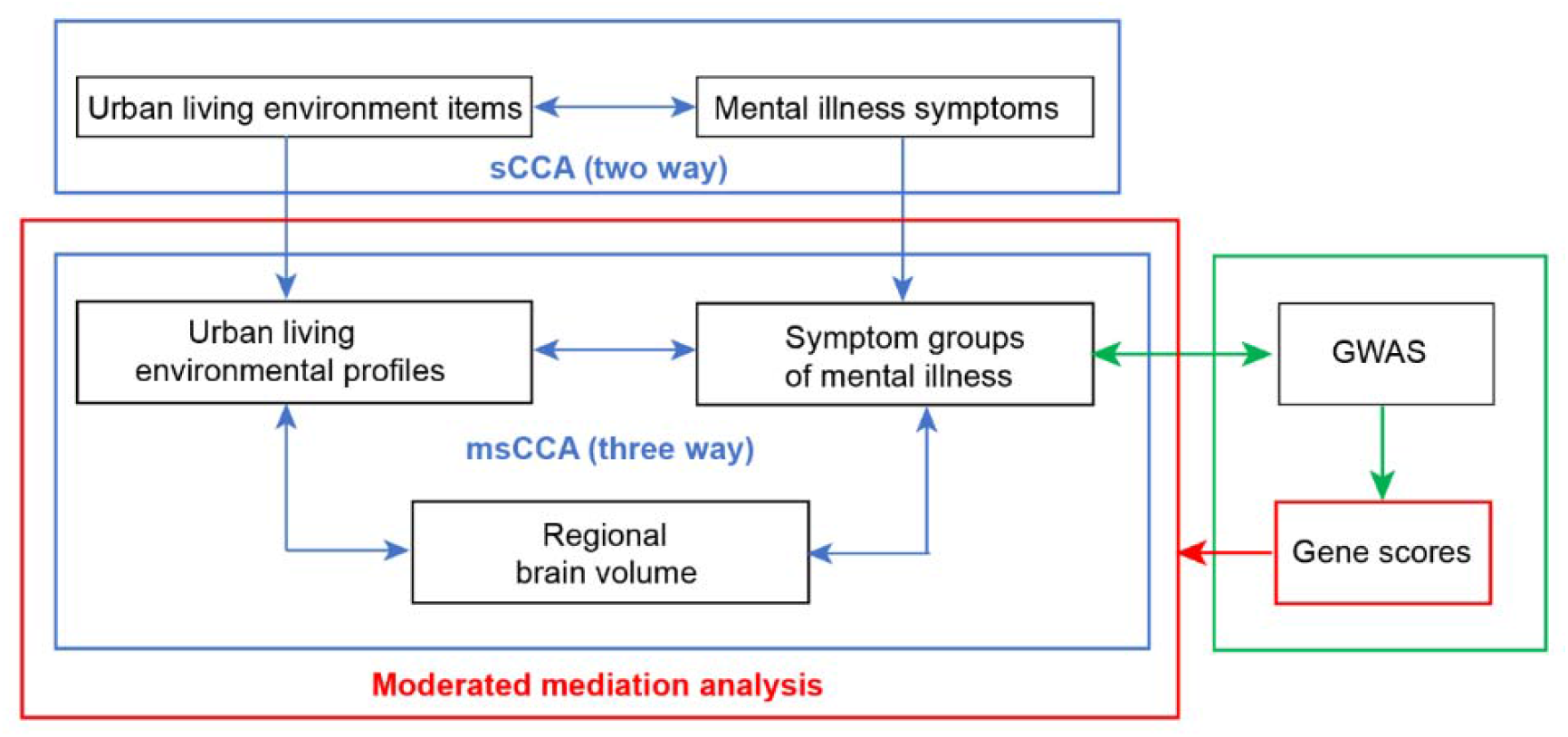
Characterization of the relation of urban living environment profile, regional brain volume and mental illness symptoms, and its moderation by genes. In 141,325 participants without neuroimaging data (UKB-nonNI), we identified environmental profiles correlated to symptom groups of emotional problems by splitting our data in training and test datasets, and applying bootstrapping with replacement and random resampling. Next, we carried out genome-wide association analysis (GWAS) analyses of the symptom groups identified in 85,348 participants with complete genomic, environment and mental health data from the UKB-nonNI dataset. The dataset with neuroimaging data (n=15,050, UKB-NI) was used for independent replication of the multivariate relation between urban living environment, genes and mental health, as well as for additional neuroimaging analyses. We analysed relations between the environmental profiles, regional brain volume and the emotional symptom groups applying msCCA. Again, we used a split design with a training and a test dataset. Using a moderated mediation analysis, we then investigated the mediation of the effect of the environmental profiles on emotional symptom groups by regional brain volume and its genetic moderation in 8,726 participants with complete genomic, environment, brain volume and mental health data in UKB-NI.

## Results

### Overview of analyses

Our analyses were carried out in participants of UK Biobank, a large and predominantly urban population-based cohort. A subset of 156,375 participants that were assessed for both, 21 mental symptoms measures as well as 128 environmental variables linked to their home address. These environmental variables included air and sound pollution, traffic, greenspace and coastal proximity, socioeconomic indices of multiple deprivation (IMD), building class, destination accessibility, land use density, terrain, normalized difference vegetation index (NDVI) and street network accessibility. The participants from UK Biobank -Biobank Urban Morphometric Platform (UK-BUMP), with complete urban living environmental data and mental health data (n=156,375) were divided into datasets without neuroimaging data (n= 141,325, UKB-nonNI) and with neuroimaging data (n=15,050, UKB-NI). At the time of our analyses, brain neuroimaging was ascertained in 42,796 participants, of which 15,050 had complete neuroimaging, mental health and environmental assessments (Supplementary methods).

We apply a training and test data split design to investigate the relation of urban living environment with symptoms of mental illness using sparse canonical correlation analysis (sCCA), a multivariate analysis technique to determine multivariate associations between two or more sets of variables. First, we characterise the environmental profiles associated with groups of mental health symptoms using a classical (binary) sCCA. Following a GWAS analysis, we infer a set of genes scores associated with these symptoms groups. Next, using a multi-view sparse CCA (msCCA), we identify the regional brain volumes jointly associated with environment and mental health. We then describe the directionality of this relationship using a moderated mediation analysis. Throughout the analysis we were careful to retain a strict training-test separation to avoid bias. Demographic information on the specific statistical analysis is shown in Table 1. Schematic summaries are shown in Figure 1 and Supplementary Figure 1.

**Table 1.**
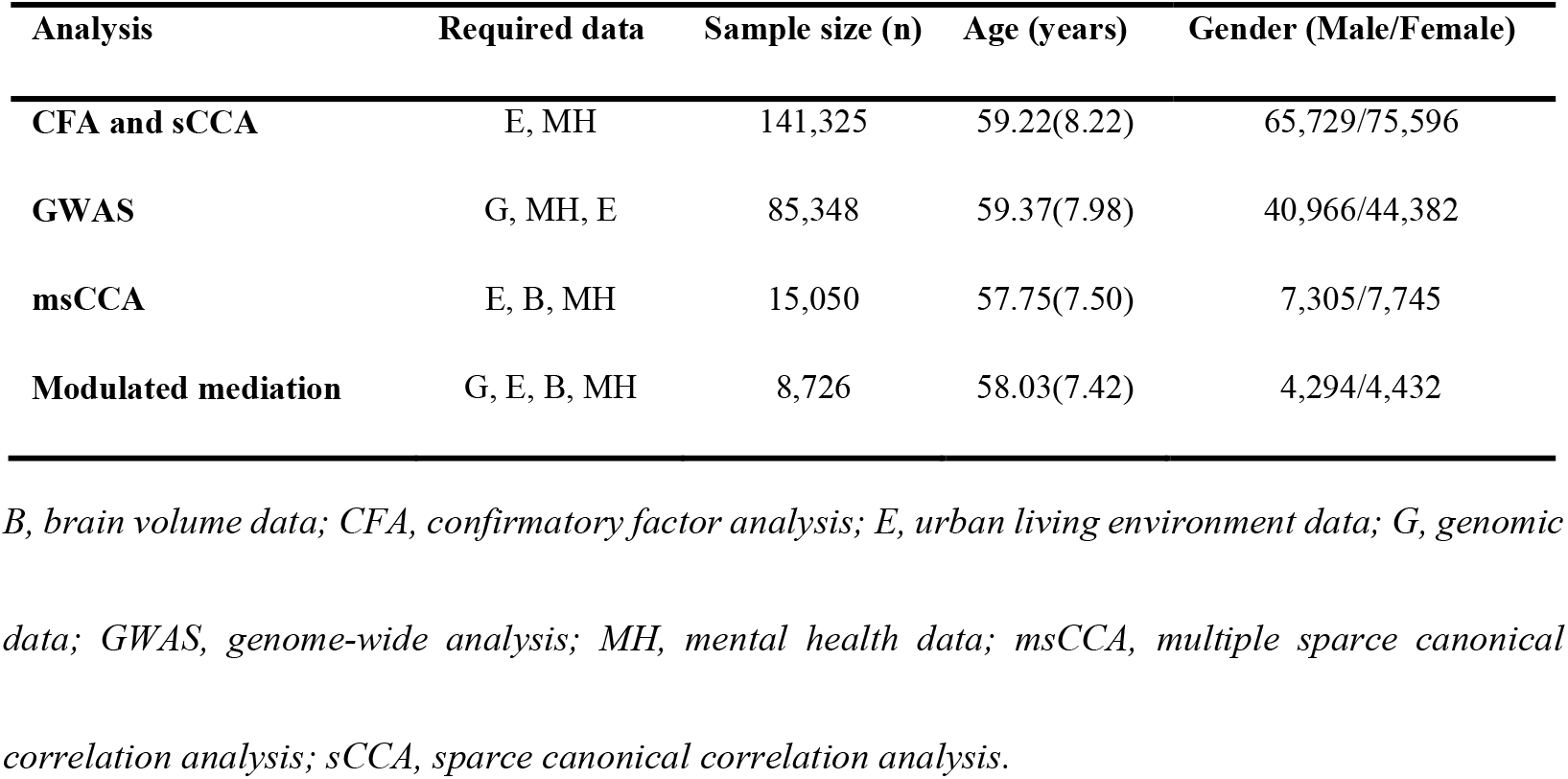
Demographics of participants used in specific statistical analysis.

| Analysis | Required data | Sample size (n) | Age (years) | Gender (Male/Female) |
| --- | --- | --- | --- | --- |
| **CFA and sCCA** | E, MH | 141,325 | 59.22(8.22) | 65,729/75,596 |
| **GWAS** | G, MH, E | 85,348 | 59.37(7.98) | 40,966/44,382 |
| **msCCA** | E, B, MH | 15,050 | 57.75(7.50) | 7,305/7,745 |
| **Modulated mediation** | G, E, B, MH | 8,726 | 58.03(7.42) | 4,294/4,432 |

*B, brain volume data; CFA, confirmatory factor analysis; E, urban living environment data; G, genomic data; GWAS, genome-wide analysis; MH, mental health data; msCCA, multiple sparse canonical correlation analysis; sCCA, sparse canonical correlation analysis.*

### Correlation of urban living environment with symptoms of mental illness

A total of 53 urban living environment catgories including 128 items were included in this study. In the 53 categories, there were 34 categories having one independent item. In the remaining 19 categories, given that some items of each category examined similar aspects of urban environment, ten fold cross-validation confirmatory factor analysis (CFA) was performed to collapse the available information into 19 latent urban living environmental categories using the R package *lavaan* (https://cran.r-project.org/web/packages/lavaan)^18^. Ten-fold cross-validation splits were performed to ensure unbiased estimates of generalizability throughout the analytic pipeline and to optimize the CFA models. To investigate the relation of urban living environment with symptoms of mental illness, we used sCCA regression to link 53 independent urban living environment categories and 21 prospective symptoms of mental illness (see Online Methods). To avoid overestimating the variance shared between urban living environment and symptoms of mental illness, we used a split data analysis design, which allows us to estimate effect sizes in an unbiased way. We carried out model selection in a training dataset of 90% of the data (n = 127,134), and model validation in the testing dataset of the remaining 10% (n = 14,132) in the 141,326 participants of the UKB-nonNI dataset. To enhance stability, we resampled the data and retained only variables that contributed to the model in 90% of resamples (Methods and Supplementary Figure 1)^19^.

Using msCCA regression, we found a significant relationship between urban living environment profiles and a subset of five symptoms of mental illness in the training dataset (*r*=0.22, *P*_permutation_<0.001), explaining 4.84% of the variance. The model was also significant when applied to the test dataset (*r* = 0.19, *P*_permutation_<0.001, *P*_FDR_<0.001) explaining 3.61% of the variance between environment profiles and symptoms groups of mental illness (Figure 1). These symptoms of mental illness consist of a group of five self-reported symptoms, namely frequency of unenthusiasm, frequency of tiredness, loneliness, frequency of depressed mood and fed-up feelings (Figure 2), which we summarised as the affective symptom group. This symptom group is positively correlated with IMD score, air and noise pollution, measures of street accessibility and neighbourhood walkability (street radial and centrality), traffic and density of urban infrastructures (factories, retails, offices and community services), while being negatively correlated with percentage of domestic garden, natural environment and greenspace, and distance to urban facilities (community services, factories, emergency, education, food stores, community and healthcare) (Supplementary Table 5 and Figure 2). We assessed robustness in two ways, first we used bootstrapping to resample the training data (with replacement) 1000 times, each containing 10% to 150% of the training dataset in 10% increments. Then, we performed a second validation by drawing 1000 resamples of the training dataset, each containing 90% of the dataset (Supplementary Table 6 and Figure 2). No specific gender effect was detected (Supplementary Results and Supplementary Table 7). Our results indicate that the affective symptom group is positively correlated with an environment profile dominated by high degrees of social deprivation and air pollution, and to a lesser extent traffic and short distance to infrastructural facilities. Other properties of urbanicity, in particular various forms of green space and social infrastructure appear to be protective in this model.

**Figure 2.**
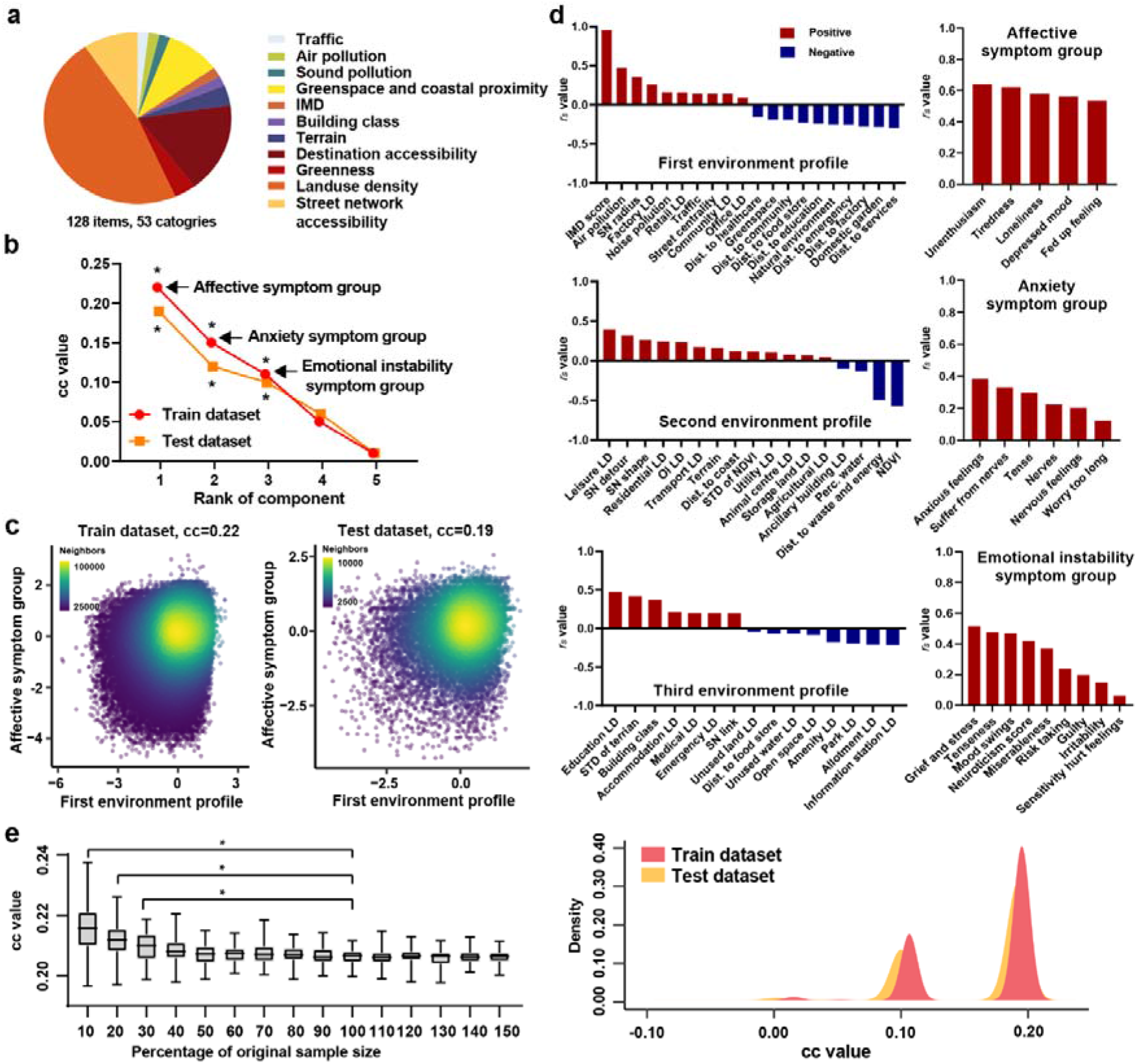
Multivariate relation between urban living environment profile and symptoms of mental illness. a. A total of 128 items corresponding to 53 categories of urban living environment are included; b. The sCCA-regression model linking urban living environment to symptoms of mental illness identified three significant correlates in train datasets (red dot), including affective symptoms group (r=0.22, P_permutation_<0.001), anxiety symptoms group (r=0.15, P_permutation_<0.001) and emotional instability symptoms group (r=0.12, P_permutation_<0.001). These results were still significant in test datasets of affective (r=0.19, P_permutation_<0.001), anxiety (r=0.12, P_permutation_<0.001) and emotional instability symptoms group (r=0.10, P_permutation_<0.001). c. An example of correlation map between urban environment profile and affective symptoms group in train dataset (r=0.22, P_permutation_<0.001) and test dataset (r=0.19, P_permutation_<0.001). d. In the first (top), second (medium) and third (bottom) correlates, urban living environment profiles contributing to this relationship were shown on the left, symptoms group of mental illness contributing to this relationship were shown on the right. The bars demonstrated the structure coefficient value (r_s_) of each variable in sCCA-regression analysis. e. Robustness assessment. Left: We used bootstrapping to resample the training data (with replacement) 1000 times, each containing 10% to 150% of the training dataset in 10% increments Stability in correlation coefficient after about 30% of the sample size were observed; Right: To estimate of the stability of the findings across subsamples, we resampled the same proportion 90% of original sample size as train dataset for 1000 times, reran the sCCA algorithm and calculated the correlation between the resulting feature in the remaining 10% test dataset. CC value, canonical correlation coefficient; IMD, Index of Multiple Deprivation; NDVI, Normalized difference vegetation index; r_s_, structure coefficient value; STD, standard deviation; UE, urban living environment.

After determining the significance of the first canonical correlate, we removed the effect of the first set of canonical vectors by projection deflation, as we have done previously^19^. This approach is more appropriate for regularised CCA than standard deflation methods more commonly used in the field^20^ because it ensures that the predictions for successive components are orthogonal (i.e. uncorrelated). This is also important to keep in mind in order to correctly interpret our findings (e.g. the GWAS analysis we report below) because the second canonical variables relate to covariance between datasets that is orthogonal to the first set of canonical variables, as is also the case in standard CCA. In other words, the second CCA component explains covariance over and above what is explained by the first component. We then repeated the analysis to investigate the presence of a second canonical correlation between the remaining urban living environment and mental health variables. Here, we identified another mental illness symptoms consisting of six self-report measures, including worrying too long, anxious feelings, nervous feelings, nerves, tense and suffering from nerves (Figure 2), summarised as the anxiety symptom group, which was significantly associated with the environmental profile in the training dataset (*r* = 0.15, *P*_permutation_<0.001). The test sample correlation is also significant (*r* = 0.12, *P*_permutation_<0.001, *P*_FDR_<0.001), explaining 1.44% of the variance between environmental profile and anxiety symptom group. This symptom group was positively correlated with measures of urban morphology, including density of leisure places, street accessibility and neighbourhood walkability (street network detour and shape) (Supplementary methods and Supplementary Table 2-3), mean terrain, Euclidean distance to coast, variation of NDVI and density of mixed urban infrastructure (residential, transport, utility, animal centre, storage land and agriculture, et.ac) while being negatively correlated with mean NDVI, distance to waste and energy as well as percentage of water (Supplementary Table 5 and Figure 2). The second environmental correlate captures a different urban profile, one that is dominated by green spaces and long distances to waste and energy facilities as well as presence of water, all of which are inversely correlated (protective) to symptoms of anxiety. The anxiety symptom group is also positively correlated with signs of denser urban build-up, such as density of streets and leisure places as well as urban regions with mixed residential, commercial and industrial use.

We removed the effects of the second canonical correlate and investigated the presence of a third canonical correlation between the remaining urban living environment and symptoms of mental illness. Here, the correlation coefficient between two canonical variables was 0.12 in the training dataset (*P*_permutation_<0.001) and 0.10 in the test dataset (*P*_permutation_<0.001, *P*_FDR_<0.001), explaining variance of 1.44% and 1.00%, respectively. The canonical mental health correlate consists of nine symptoms including feeling guilty, frequency of tenseness, miserableness, mood swings, neuroticism score, risk taking, irritability and sensitivity, hurt feelings, illness, injury, bereavement or stress in last 2 years (Figure 2), which we termed the emotional instability symptom group. This symptom group is positively correlated with density of education facilities, variation of terrain, building class (flats in highrisers, terraced houses etc.), measures of street accessibility and neighbourhood walkability (street network link characteristics), density of accommodation, medical and emergency facilities, while being negatively correlated with density of unused land, density of water, open space, amenity, park, allotment and information stations as well as distance to food store.

We independently replicated these correlations in the UKB-NI dataset (n=15,050) by applying the same sCCA split data (90%/10%) with a resampling threshold of 90%. The replication analysis yielded three statistically significant canonical correlates in the training and test datasets, which were identical to those of the primary analyses. In the first affective symptoms group, the canonical correlation was 0.20 in the training dataset (n=13,545; *P*_permutation_<0.001) and 0.16 in test dataset (n=1,505; *P*_permutation_<0.001, *P*_FDR_<0.001), in the second anxiety symptoms group, the correlation coefficient was 0.10 in the training dataset (*P*_permutation_<0.001) and 0.08 in the test dataset (*P*_permutation_<0.001, *P*_FDR_<0.001). In the third emotional instability symptoms group the correlation coefficient between two canonical variables was 0.06 in the training dataset (*P*_permutation_<0.001) and 0.04 in the test dataset (*P*_permutation_<0.0039, *P*_FDR_ =0.0273) (Supplementary Table 8).

### GWAS analyses of the mental health symptoms groups that related to urban environment profile

We next performed a GWAS analysis of the canonical variates of the affective, anxiety and emotional instability symptom groups in 85,348 participants with complete genetic, urban environment and mental health data in UKB-nonNI datasets (Table 1). Gene-set enrichment analysis was then performed to explore biological mechanism underlying the genes associated with the symptoms groups using TopGene^21^. To reduce dimensionality, we generated scores for each of the genes where significant SNPs were localised (see Supplementary methods). The score of each gene is calculated as the sum of the count of risk alleles multiplied by the corresponding beta value from GWAS across the index SNPs of each clump after adjusting for linkage disequilibrium (see Supplementary methods). These gene scores were then analysed for moderation of the relation of environment, regional brain volume and symptoms group of mental illness (see below).

For the affective symptom group, we found 2,983 significant associations with SNPs at Bonferroni *Pc*<0.05 (uncorrected *P*<0.05/139,187,27/3=1.20 × 10^−9^, Supplementary Table 9), located in 43 genes. The lambda genomic control inflation factor was 1.0195 and the intercept value in linkage disequilibrium score regression (LDSC) was 1.011, indicating that population stratification was minimal in this study and did not cause significant inflation to the test statistics. By far the strongest association with the affective symptom group was found in a genomic region of chromosome 17q21.3 spanning from position 43487217 to 44862162 (hg19), the site of a human supergene candidate that encodes several genes previously implicated in mental illness (Figure 3)^22^. The lead SNP was rs62062288, located in intron 6 of microtubule associated protein tau (*MAPT*) gene of chromosome 17 (*P*=6.09×10^−15^), a gene that encodes Tau protein in neurons and has been shown to be involved in affective symptoms^23^, alcohol disorders^24^ as well as risk taking behaviour^25^. In the same region of chromosome 17q21.3, we also found strong association of the affective symptom group with Corticotrophin Release Hormone Receptor 1 (*CRHR1*, a critical regulatory gene for neuroendocrinological and behavioural stress response^26^ as well as alcohol use disorders^27,28^). The remaining top associated genes were also encoded in this region. They include *ARL17B* (ADP ribosylation factor-like GTPase17B, potentially regulating G-protein signalling^29^), *KANSL1* (KAT8 regulatory NSL complex subunit1, part of a histone acetyltransferase complex, an epigenetic regulator^30^), and *MAPT-AS1* (MAPT antisense RNA 1, involved in progression of tau pathology in neurodegeneration^31^), as well as a pseudogene *RP11-259G18* (Figure 3 and Supplementary Table 9). Additional associations were found on genomic region of chromosome 18q21.2, at the *DCC* gene locus (DCC Netrin 1 Receptor, regulates synaptic plasticity^32^ and neuronal migration^33^) and *TCF4* gene locus (transcription factor 4, involved in neural differentiation^34^), chromosome 14q24.1 (*DCAF5* and *EXD2* gene locus) and chromosome 3q22 (*STAG1* and a pseudogene *RP11-102M11.2*). Bioinformatics analyses of the 43 genes using TopGene revealed enrichment of neuronal differentiation, regulation of axogenesis and, importantly, CRH-binding and activity (Supplementary Table 12). We then calculated 43 genes scores. The relationship between urban living environment profile and affective symptom group have shown different vulnerabilies depending on the genetic background. For example, we found that participants with lower *CRHR1* gene scores demonstrate smaller correlation of urban environment profile with the affective symptom group compared to those with higher *CRHR1* genes scores (Figure 3).

**Figure 3.**
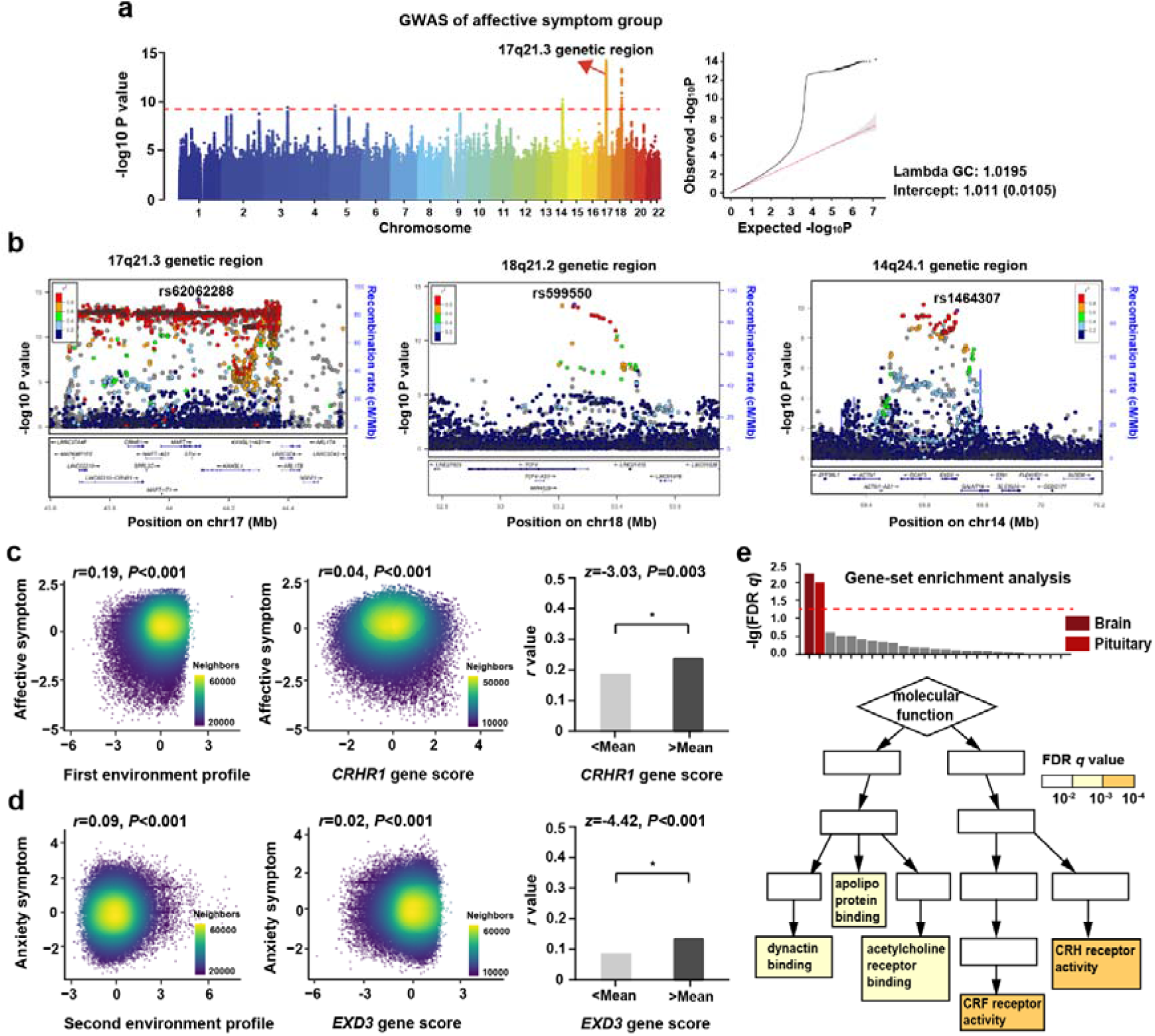
Associations of genomics, urban living environment profiles and mental health symptoms. a. Left. Manhattan plot of GWAS analysis. GWAS of the affective symptom group identifies 2,983 significantly associated SNPs. The strongest association with the affective symptom group was found in a genomic region of chromosome 17q21.3 spanning from position 43487217 to 44862162 (hg19) containing a potential supergene that encodes several genes previously implicated in mental illness. An intron variant rs62062288 located in MAPT gene of chromosome 17 (P=6.09×10^−15^) was the lead SNP; Right. Quantile-quantile plot of GWAS analysis. The lambda genomic control inflation factor of 1.0195 and intercept value in LDSC of 1.011, which indicate that population stratification was minimal in this study to cause significant inflation to the test statistics. b. Locus zoom plots of significant genomic region of 17q21.3 (left), 18q21.2 (medium) and 14q24.1 (right) in GWAS analysis of affective symptom group. The purple dots demonstrate the lead SNPs of each genomic region. c-d. Two examples of the affective symptom group influenced by both genomics and urban living environment profile. c. Left. The first urban living environment profile showed significant positive correlation with the affective symptom group; Medium. Participants with higher CRHR1 gene scores show more severe affective symptoms compared to those with lower CRHR1 gene scores; Right: Participants with lower CRHR1 gene scores demonstrate smaller correlation of urban environment profile with the affective symptom group compared to those with higher CRHR1 genes scores. d. Left. The second urban living environment profile showed significant positive correlation with the anxiety symptom group; Medium. Participants with higher EXD3 gene scores show more severe anxiety symptoms compared to those with lower EXD3 gene scores; Right: Participants with lower EXD3 gene scores demonstrate smaller correlation of urban environment profile with the anxiety symptom group compared to those with higher EXD3 genes scores. e. Gene-set enrichment analysis of affective symptom group-associated 43 genes. Top: Tissue-specific expression analysis showed affective symptom group associated 43 genes were significantly overexpressed in brain and pituitary tissues. Bottom: In the 43 genes associated with the symptom group, we found over-representation in the molecular function of CRH receptor activity (FDR qc = 4.37 × 10^−4^) and CRF receptor activity (FDR qc = 4.37 × 10^−4^); CRH, corticotropin-releasing hormone; CRF, corticotropin-releasing factor; GWAS, genome-wide association analysis; lambda GC, lambda genomic control inflation factor.

We found significant associations of the anxiety symptom group with 29 SNPs covering 11 genes at Bonferroni *Pc*<0.05 (Supplementary Table 10 and Supplementary Figure 2). The drop in genome-wide significant hits compared to the affective symptom group GWAS is likely caused by the decreased covariance of the second CCA correlate, the anxiety symptom group, following deflation of the correlates of the first CCA correlate, the affective symptom group, as described above. The lead SNP associated with the anxiety symptom group was rs77641763 located in the intron 15 of Exonuclease 3’-5’ Domain Containing 3 (*EXD3*) gene of chromosome 9 (*P*=4.08×10^−10^). *EXD3*, involved in nucleic acid binding and widely expressed in the brain, has been associated with a group of anxiety, phobic and dissociative disorders^35^. The other top significant genes include *NOLC1* (Nucleolar And Coiled-Body Phosphoprotein 1), a chaperone for shuttling between the nucleolus and cytoplasm^36,37^, *ELOVL3* (Elongation of very long chain fatty acids protein 3, provides precursors for synthesis of sphingolipids and ceramides^38^), *LBX1-AS1* (Ladybird homeobox 1-antisense RNA 1, involved in neuronal determination processes^39^), *PITX3* (Paired Like Homeodomain 3, regulates differentiation and maintenance of midbrain dopamine neurons during development^40^), *GBF1* (Golgi Brefeldin A Resistant Guanine Nucleotide Exchange Factor 1, involved in axonal neuropathy^41^), *CNNM2* (Cyclin And CBS Domain Divalent Metal Cation Transport Mediator 2, linked to neurodevelopmental impairments^42^) and *NT5C2* (cytosolic 5′-nucleotidase II, regulating AMPK signalling and protein translation during early neurodevelopment^43^) (Supplementary Table 10). The 11 genes associated with the anxiety symptom group were enriched for small nucleolar ribonucleoprotein (snoRNP) complex binding involved in serotonin receptor regulation (Supplementary Table 12). And participants with lower *EXD3* gene scores demonstrate smaller correlation of urban environment profile with the anxiety symptom group compared to those with higher *EXD3* genes scores (Figure 3).

In the emotional instability symptom group, there were 6 significant genetic associations at Bonferroni *Pc*<0.05 (Supplementary Table 11 and Supplementary Figure 2). The lead SNP was rs77786116 located in Intraflagellar Transport 74 (*IFT74*) gene of chromosome 9 (*P*=4.16×10^−10^). *ITF74* is a critical factor in neuronal migration, a cause for Bardet-Biedl syndrome and Joubert syndrome^44,45^ and is associated with paranoid schizophrenia^46^. The other significant genes include *TMPO* (thymopoietin, involves in neuron proliferation), *LDHC* (lactate dehydrogenase C, involved in anaerobic and aerobic glycolysis), *SLC9A7P1* (solute carrier family 9 member 7 pseudogene 1). Together, they are enriched for cerebellar granular layer development process (Supplementary Table 12).

We independently replicated the SNPs that in the discovery GWAS were significantly associated with symptom groups of mental illness (UKB-nonNI dataset) in a dataset of 8,726 participants of UKB-NI. The significance threshold was Bonferroni *Pc*<0.05 (uncorrected *P*<0.05/3018, the numbers of all significant SNPs from discovery GWAS analysis). Of these 3018 significant SNPs from GWAS analysis of mental illness symptom groups, we replicated in the independent UKB-NI dataset 2034 SNPs associated with the affective symptom group, 18 SNPs associated with the anxiety symptom group and 3 SNPs associated with the emotional instability symptom group (Supplementary Table 13-15). We then calculated the corresponding genes scores as before and validated the associations between genes scores and symptoms group of mental illness in the UKB-NI dataset. Of 43 genes scores associated with affective symptom group in the discovery analysis, we replicated 36 genes in the replication analysis, the top ten associated genes including *MAPT, CRHR1, DCC, DND1P1, KANSL1, PLEKHM1, RP11-363J20.1:EXD2, STAG1, C5orf17 and DCAF5*. Of 11 genes scores in the anxiety symptom group, we replicated 6 genes, including *EXD3, LBX1-AS1, NT5C2, GBF1, NOLC1* and *CNNM2*. Of 6 genes scores associated with emotional instability symptom, we replicated 3 genes including *LDHC, IFT74* and *TMPO* (Supplementary Table 15).

### Brain volume changes underlying the associations between urban living environment and symptom groups of mental illness

To further investigate the neurobiological mechanisms underlying the associations of urban living environment with mental symptoms groups, we carried out msCCA on the urban living environment profiles, brain volume measures and symptoms group of mental illness. This analysis was conducted in an independent dataset of 15,050 UKB participants, split in a training dataset (90%) and a test dataset (10%). We found 13 regional brain volumes significantly associated with the first urban environmental profile and the affective symptom group (*r*=0.052, *P*_permutaiton_=0.01 in training dataset; *r*=0.046, *P*_permutaiton_=0.01 in test dataset). These brain volumes include the left amygdala and right ventral striatum, right frontal pole, right occipital fusiform gyrus, as well as bilateral cerebellar lobules VIIIa and VIIb, right posterior cerebellum crus I and II (Figure 4 and Supplementary Table 17). The first urban environment profile was negatively correlated with brain volume in these areas and positively correlated with affective symptoms group. We also found 11 regional brain volumes significantly associated with the second urban environmental profile and the anxiety symptom group (*r=*0.045, *P*_*permutation*_=0.006 in training dataset; *r=*0.045, *P*_*permutation*_=0.007 in test dataset) (Supplementary Table 17). These brain volumes include the inferior frontal regions and the right amygdala, as well as the bilateral cerebellar lobules VIIIa and VIIb, posterior cerebellum crus I, right cerebellar lobule V and left lobule VI (Figure 4 and Supplementary Table 17). Finally, there were 13 regional brain volumes, including bilateral frontal pole, amygdala, precentral gyrus, insular and left lateral occipital cortex, associated with the third urban environment profile and the emotional instability symptom group (*r=*0.057, *P*_*permutation*_<0.001 in the training dataset; *r=*0.060, *P*_*permutation*_<0.001 in test dataset) (Figure 4 and Supplementary Table 17).

**Figure 4.**
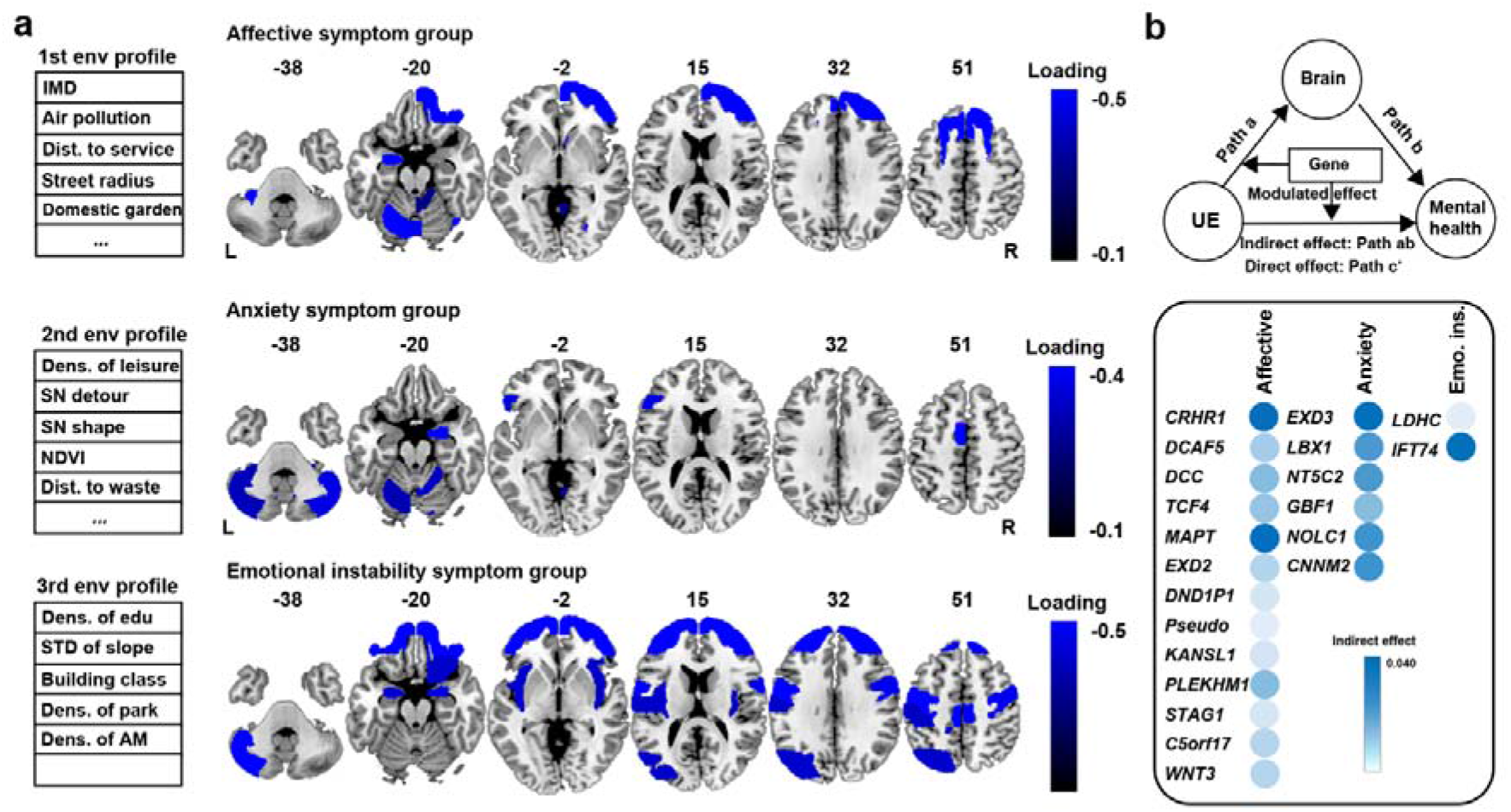
Three-way msCCA regression between urban living environmental profile, regional brain volume and mental illness symptom group, and moderated mediation analysis of gene scores. a. Left. Urban living environment variables contributing to the three way corelates are shown on the left. Righ. Regional brain volume associated with urban living environment profile and affective (top), anxiety (medium) and emotional instability (down) symptom groups, respectively, in both training and test datasets. Negative loading was shown in blue. b. Top. A schematic diagram of moderated mediation analysis between genomics, urban living environment profile, brain and mental illness symptom group. Bottom. Columns show an indirect effect in the mediation analysis between environmental profile, brain volume and mental illness symptom groups, which were moderated by each gene score. We found that the CRHR1 gene score (EME=5.01%), MAPT gene score (EME=3.62%), TCF4 gene score (EME=1.62%) and DCC gene score (EME=1.96%) moderate the mediation pathway from environmental profile to brain correlate to the affective symptom group. The EXD3 gene moderates the mediation pathway from environmental profile to brain correlate to the anxiety symptom group (EME=3.65%). IFT74 gene score moderates the mediation pathway from environment profile to brain correlate to emotional instability symptom group (EME=2.86%). AM, allotment. EME, explained mediation effect; IMD, Index of Multiple Deprivation; NDVI, normalized difference vegetation index; SN, street network measures; STD, standard deviation.

### Moderated mediation analysis between urban living environment profile, brain volume and mental illness symptoms groups modulated by genomics

To test whether the relation of urban living environment with symptoms group of mental illness is mediated by brain volume and moderated by genetics, we independently performed moderated mediation analysis in each replicated gene score (moderating variable), three urban living environment profiles (independent variable), three brain volume correlates (mediated variable) and three symptoms groups of mental illness (dependent variable) in 8,726 participants with complete data. Explained mediation effect (EME) were reported for moderated mediation analyses. Thus, a total of 21 moderated mediations analysis were tested (13 gene scores of the affective symptom group, 6 gene scores of the anxiety symptom group and 2 gene scores of the emotional instability symptom group). Of the 13 replicated gene scores in the affective symptom group, we found that the *CRHR1* gene score (EME=5.01%), *MAPT* gene score (EME=3.62%), *TCF4* gene score (EME=1.62%) and *DCC* gene score (EME=1.96%) moderate the mediation pathway from environment profile to brain correlate to affective symptom group. For example, participants with higher *CRHR1* genetic risk living in areas of more urban environment exposure had lower brain volume and demonstrated more severe affective symptoms (Figure 4). We found moderation of the mediation pathway of the anxiety symptom group by the *EXD3* gene score (EME=3.65%) and of the emotional instability symptom group by the *IFT74* gene score (EME=2.86%) (Figure 4).

## Discussion

We describe how the urban living environment affects brain and mental illness by identifying environmental profiles derived from a comprehensive set of measures of deprivation, pollution, land use, and infrastructure that are correlated with distinct groups of affective, anxiety and emotional instability symptoms, mediated by reductions in regional brain volume and moderated by genes involved in pertinent biological pathways.

Our approach and findings are novel, as they characterise specific multimodal environmental profiles in an integrated way, while enabling a qualitative and quantitative assessment of each factor of the profile. This is an advance beyond the isolated assessment of individual environmental factors, as has previously been the norm^9,47^. As a consequence, we are able to explain a greater degree of variance of integrated environmental factors (3.61%) than comparable studies measuring individual environmental factors, such as night-time light (2.56%), built-up areas (1.21%) and NDVI (1.00%)^8^, alone. Furthermore, the integrated approach enables us to assess the role of each individual environmental factor in a context that is relevant for mental illness. In another innovation, we describe how the effect of environmental profiles on symptoms groups of mental illness is mediated by regional brain volume and moderated by genetic factors.

By providing evidence for the brain-related correlates of environmental adversity and their consequences for mental health, we are broadening the evidence required for responsible urban and development planning. Furthermore, we are enabling the development of neurobehavioural interventions that may convey adaptive coping skills for environmental adversity by targeting specific brain mechanisms, for example through neurofeedback-guided virtual reality sessions^48,49^. The identification of genetic moderators points towards biological pathways that might underlie the observed relation between environment, brain and behaviour. It may also enable the selection of individuals that are more sensitive to environmental adversity, and thus might be more likely to benefit from targeted interventions.

Specifically, our analyses reveal the first environmental profile dominated by high degrees of deprivation and air pollution, and to a lesser extent traffic, short distance to infrastructural facilities and lack of green space. This environmental profile evokes the image of a poor, dense inner city neighbourhood. It is correlated with increased affective symptoms, in particular high levels of unenthusiasm, tiredness, loneliness, as well as depressed mood and feelings of being fed-up. The correlation is mediated by volume reductions in brain regions linked to emotional processing, such as the left amygdala^50^, regions linked to reward processing and exploration, such as the right ventral striatum^51^ and the right frontal pole^52^, as well as other brain areas implicated in affective processing, such as several cerebellar regions^53^ and the right occipital fusiform gyrus^54^. While these findings point towards plausible mediators of a stressful environment on affective symptoms, they also suggest the presence of different underlying neurobehavioural mechanisms, a possibility that is supported by the findings of our genetic analysis. Here we identified different moderators that may influence distinct brain mechanisms underlying the affective symptoms identified: First and foremost, *CRHR1*, which is also expressed in the amygdala^55^, is a critical regulator of both the hypothalamic as well as the behavioural extrahypothalamic stress response. In addition, the environmental effect is moderated by genes regulating brain structure, including *MAPT*, involved in neurodegeneration^56^, *TCF4*, inducing neural differentiation^57^ and *DCC*, an adhesion molecule that guides axon growth^58^. Apart from these moderators, we identified several genes that were associated with the affective component and were involved in relevant neural mechanisms, including G-protein signalling (*ARL17B*)^59^, epigenetic regulation (*KANSL1*)^60^ and others. These moderating genes are located in two genomic loci of chromosome 17q21.3 and 18q21.2. Notably, the chromosome 17q21.3 genomic locus is the site of a human supergene candidate, a cluster of tightly linked functional genetic elements spanning approximately 900 kb that control balanced phenotypes and are inherited as a unit^61^. Haplotypes of this cluster have been associated with brain morphology and different cognitive and behavioural traits, including depressive behaviour, neuroticism and risk taking behaviour^62,63^. The 18q21.2 region at the transcription factor 4 gene *TCF4* and netrin 1 receptor gene *DCC* featured the most pleiotropic association with eight psychiatric disorders^64^, and has been previously associated with both depression and neuroticism^65,66^. The product of *DCC* plays a critical role in guiding axonal growth during neurodevelopment and serves as a master regulator of midline crossing and white matter projections^67^.

The second environmental profile captures a different urban profile, one that is dominated by green spaces and long distances to waste and energy facilities as well as presence of water, all of which are inversely correlated (protective) to symptoms of anxiety. These symptoms are ‘worry too long, anxious feelings, nervous feelings, nerves, tense and suffer from nerves’. The anxiety symptoms are also positively correlated with signs of denser urban build-up, such as density of streets and leisure places as well as urban regions with mixed residential, commercial and industrial use. Together, these correlations point to an important role of green spaces and a less dense and more generous land use in protecting against symptoms of anxiety, thus extending previous findings relating urban green spaces to mental health^8^. The relation between environmental profiles and anxiety symptoms are mediated by volume reductions in the inferior frontal regions, the right amygdala and cerebellar regions. They are moderated by variations in the *EXD3* gene, involved in nucleic acid binding and widely expressed in the brain. *EXD3* has previously been associated with anxiety, phobic and dissociative disorders^35^. While *EXD3* has a low tissue specificity, its expression in the brain is highest in frontal cortical brain areas. Similar to the second environmental profile, the third environmental profile shows positive correlations of measures of density of land use and urban infrastructure with a group of symptoms best summarised as emotional instability, which include feeling guilty, frequency of tenseness, miserableness, mood swings, neuroticism score, risk taking, irritability and sensitivity, hurt feelings, illness, injury, bereavement or stress in last 2 years. These symptoms were mediated by frontal pole, amygdala, precentral gyrus, insular cortex and cerebellum, and moderated by *ITF74*, a critical factor in neuronal migration, a cause for Bardet-Biedl syndrome and Joubert syndrome^44,45^, which has been found associated with paranoid schizophrenia^46^.

While our findings differentiate distinct groups of environmentally-related symptoms of mental illness, the overarching role of social deprivation^68^ and urban density^69^ as a risk factor is evident. Conversely, we found green spaces^9,10^ and generous land use^70^ to be protective factors against affective and anxiety symptoms. These environmental factors may engage brain mechanisms related to behavioural inhibition (frontal cortex), reward processing (ventral striatum) and emotional processing (amygdala) that may be moderated by biological pathways involved in stress response, neuronal plasticity, epigenetic, transcriptional and neurotransmitter regulation. Interestingly, the posterior cerebellar lobule VIIb and VIIIa, posterior cerebellar crus I are among the brain mediators of environmental effects on symptoms of mental illness, supporting a role for the cerebellum in coordinating and regulating not only voluntary movements, but also cognitive and affective perceptions of the environment^8,71^.

Our data do not enable the characterisation of individual biological pathways that mediate defined environmental adversity. To carry out such mechanistic investigation and to identify biomarkers for risk and resilience, a more deeply phenotyped dataset with neurobehavioural characterisation is required. Our findings generate hypotheses, that may be tested in well characterised samples of a much smaller size. Another important question raised by this study is the generalizability of its results beyond industrialised high income countries to low and medium income countries, where socioeconomic conditions between rural and urban dwellers may be different compared to Europe. Further studies validating our results in a global mental health context are required.

By providing first evidence for comprehensive environmental profiles that affect distinct groups of symptoms of mental illness, our results lay the ground work for an exact characterisation of biological mechanisms underlying complex, real-life environmental adversity. The quantification of the relative contribution of each environmental factor to brain and symptoms of mental illness and their interplay in a living environment are novelties of this study that will aid in targeting and prioritizing important decisions for planning and public health interventions.

## Material and methods

### Participants

#### UK Biobank Project

UK Biobank (UKBB) is a population-based cohort including 502,616 participants living in United Kingdom, where over 40,000 participants have obtained neuroimaging scans (at the time of our analyses datasets of 42,796 participants were available). Individuals were invited to the study if they were registered with the National Health Service and if they lived within a 35 km radius of one of the 22 assessment centres located across the UK at the time of recruitment, which took place between 2007 and 2010. Baseline assessments included genomics, physical and social exposures, sociodemographic, lifestyle, occupational, psychosocial and environmental measures. Informed consent was obtained from all UKBB participants. Ethical procedures are controlled by a dedicated Ethics and Guidance Council (http://www.ukbiobank.ac.uk/ethics) that has developed with UKBB an Ethics and Governance Framework (http://www.ukbiobank.ac.uk/wp-content/uploads/2011/05/EGF20082.pdf), with IRB approval also obtained from the North West Multi-center Research Ethics Committee. The data collected at baseline was used in this study. Demographic information of each statistical analysis was shown in Table 1.

### Data collection

#### Urban living environment data

Indices of multiple deprivation (IMD), traffic, residential pollution, greenspace and coastal proximity as well as urban morphometric measures were used to measure the urban living social and physical environment around participants available in the category ‘local enviroment’ in UK Biobank (data-field 113). A total of 53 categories including 128 urban living environment items were included in the urban living environment data. The detailed items and categories used are shown in Supplementary Table 1 and 2. To exclude items with extremely skewed data distribution, we used the function *nearZeroVar* from the *caret* R package^72^ and no items excluded. In the 128 items, we calculated the median absolute deviation (MAD) and removed values larger than 4 MAD in each environment item. For further analyses, we used 216,341 participants with complete 128 environmental items.

#### Indices of Multiple Deprivation

Indices of Multiple Deprivation (IMD) scores were used to classify the relative deprivation (a measure of poverty) in British local councils which published by UK government (https://www.gov.uk/government/collections/english-indices-of-deprivation). IMD scores were calculated separately in England (EIMD), Scotland (SIMD) and Wales (WIMD), because multiple different components of deprivation are weighted with different strengths and compiled into a single score of deprivation. The EIMD score is composed of seven domain indices, including income deprivation (Income subdomain, Income Deprivation Affecting Children Index and Older People Index), employment deprivation, health deprivation and disability, education, skills and training deprivation (children and young people subdomain and adults’ skills subdomain), barriers to housing and services (wider and geographical barriers subdomain), living environment deprivation (indoors and outdoors subdomain) and crime. The SIMD score is composed of seven domain indices, including crime (only from 2006), current income, education, skills and training, employment, geographic access, health and housing. The WIMD score is composed of eight domain indices for income, employment, health, education, access to services, community safety, physical environment and housing.

#### Traffic

Traffic consists of seven items: (1) Close to major road; (2) Inverse distance to the nearest major road; (3) Inverse distance to the nearest road; (4) Sum of road length of major roads within 100m; (5) Total traffic load on major roads; (6) Traffic intensity on the nearest major road; (7) Traffic intensity on the nearest road.

#### Residential pollution

Residential air pollution consists of five items: (1) Nitrogen dioxide air pollution from 2005 to 2010; (2) Nitrogen oxides air pollution in 2010; (3) Particulate matter 10 um air pollution in 2007 and 2010; (4) Particulate matter 2.5 um air pollution in 2010; (5) Particulate matter 2.5-10um air pollution in 2010. Residential sound pollution consists of five items: (1) Average 16-hour sound level of noise pollution; (2) Average 24-hour sound level of noise pollution; (3) Average daytime sound level of noise pollution; (4) Average evening sound level of noise pollution; (5) Average nighttime sound level of noise pollution.

#### Greenspace and coastal proximity

Greenspace and coastal proximity category contains environmental indicators relating to greenspace exposure and distance to the coast that was attributed to participants based on 300m home location buffers, including five items: (1) Natural environment percentage estimate compared to the ‘built environment’; (2) Greenspace percentage estimates; (3) Domestic garden percentage estimates; (4) Domestic water percentage estimates; (5) Distance from home location to coast.

#### Urban Morphometric Platform measures

UK Biobank Urban Morphometric Platform (UKBump) was an individual-level built environment database of health-specific urban exposures within residential street catchments of UK Biobank participants’ geocoded home address^73^. Spatial and network modelling were performed upon multiple UK-wide dataset, including AddressBase Premium data of Ordnance Survey GB, remotely sensing data, digital terrain topographical models and other datasets based on the anonymized UK Biobank participants’ geocoded home address^73^.

A total of six metrics including 104 urban environment measures from UKBump were used. The six metrics used in this study include: (1) Building class (n=3); (2) Destination accessibility (n=33); (3) Greenness (n=2); (4) Land use density (n=46); (5) Street network accessibility based on 400m home location buffers (n=18) (Supplementary Table 3); (6) Terrain (slope) (n=2).

### Genomics data

We used the imputed genomic data (Version 3) made available by UK Biobank with 487,411 individuals^74^, which was imputed from the Haplotype Reference Consortium (HRC) reference panel^75^ and a merged UK10K and 1000 Genomes phase 3 reference panels^76^. In participants-level quality control, we applied exclusion filters for participants as follows: (1) participants with a mismatch in reported sex and chromosome X imputed sex or with putative sex chromosome aneuploidy; (2) participants with genetic kinship to other participants; (3) excess heterozygosity or missing rates; (4) non-Caucasian participants; (5) without calculated genetic principal components. In SNPs-level quality control, we applied exclusion filters for SNPs as follows: (1) minor allele frequency (MAF) < 0.001; (2) imputation info quality score > 0.3. A total of 275,988 participants and 139,187,27 SNPs were finally used in the further analysis.

### Mental health measures

There are 44 mental health items in the category ‘Mental health’ in the UK Biobank that cover symptoms of affective and anxiety disorders, as well as personality (category id:100060). These items were obtained from a standardised mental health questionnaire that participants answered at the time of recruitment. Of this questionnaire 21 items were excluded because the missing rate was larger than 50% from 502,616 participants of UK Biobank. To exclude mental health items with extremely skewed data distribution, we used the function *nearZeroVar* from the *caret* R package^72^ and excluded 2 items. Finally, a total of 21 mental health items with complete data in 365,201 participants were included in the further analysis. A full list of the 21 mental health measures is shown in Supplementary Table 4.

### Neuroimaging data

In this study, neuroimaging data were acquired from one 3.0-Tesla MRI scanner from Siemens® Skyra running VD13A SP4 with a standard 32-channel radiofrequency receive head coil at UK Biobank imaging center in Cheadle Manchester. The standard parameters of a 3D MPRAGE sequence are shown in https://biobank.ctsu.ox.ac.uk/crystal/crystal/docs/brain_mri.pdf.

A total of 154 regional image-derived phenotypes (IDPs) were investigated: 111 cortical and subcortical gray matter volume (GMV) segmentations from the FAST segmentations (data-field 1101), 28 cerebellum GMV segmentations from the FAST segmentations (data-field 1101), and 15 subcortical volumes from the FIRST (data-field 1102). The detailed information are shown in https://biobank.ctsu.ox.ac.uk/crystal/crystal/docs/brain_mri.pdf. A total of 42,796 participants’ neuroimaging data were used for the present study.

### Confounding variables

Age, gender and assessment centers were adjusted as confounding covariates in the further analysis. The 21 mental health variables were firstly corrected for confounding variables and normalized as well. For neuroimaging-related analyses, total incranial volume (TIV) was additionally corrected.

## Statistical analysis

### Train and test sample split design

Participants from UK Biobank with complete urban living environmental data and mental health data (n=156,375) were divided into datasets without neuroimaging data (UKB non-NI dataset, n=141,325) and with neuroimaging data (UKB NI dataset, n=15,050). Participants without neuroimaging data (n=141,325) were divided into train and test dataset to ensure validity of the results: 90% of the participants were used as a training dataset (n=127,193) and 10% of the participants (n=14,132) were used as a test dataset for model validation. The dataset with neuroimaging data (n=15,050) was used for independent replication of the relation between urban living environment, genes and mental health, as well as for additional neuroimaging analyses. (Table 1).

### Urban living environmental variables construction

In the CFA models, ten-fold cross-validation was performed to ensure unbiased estimates of generalizability throughout the analytic pipeline and to optimize the CFA models. For each fold, 90% of participants were used to build the CFA model, and the optimized CFA model was used to calculate latent variables for the remaining 10% of participants in each environment subcategory. We used two criteria to optimize the CFA model by selecting appropriate environment measures. The first criterion was the goodness of fit of the CFA model assessed by Tucker-Lewis index (TLI), Comparative Fit Index (CFI), chi square, root mean square error of approximation (RMSEA) and standard root mean square residual (SRMR). The criteria for excellent model fit were TSI>0.95, CFI>0.95, RMSEA<0.06 and SRMR<0.08^57-59^. The second criterion was the inclusion of environment measures that best reflect different aspects of urban environment. For example, in residential noise pollution variables, we initially constructed a CFA model by including all five noise pollution measures in the training dataset. Based on factor loadings of the five noise pollution measures, we removed the ‘Average night-time sound level of noise pollution’ item with the smallest factor loading and repeated the CFA modelling. These steps were iterated until the resulting CFA model satisfied our criteria for excellent model fit in the training dataset. The factor loadings of the optimized CFA model were used to calculate the latent residential noise pollution measure in the test dataset. This process was applied into 10 folds to predict all out-of sample 19 environmental variables.

### Multivariate relation of urban living environment with mental health

To investigate the multivariate relation between urban living environment and mental health, we conducted multivariate analyses using sparse canonical correlation analysis (sCCA) implemented in MATLAB (version R2018a) based on our previous work^19^. In detail, the analysis design was carried out as follows:

1. The full dataset was randomly split into training and test datasets. The training dataset was made up of 90% of the data whilst the testing set was made up of the remaining 10%.
2. The training dataset was then randomly split into 100 resamples. Each resample was made up of nt/2 participant scans, where nt is the total number of participants in the training dataset.
3. The first stage of the msCCA-regression algorithm was then applied to each resample, with a sparsity constraint of 0.5 in each view of the data^19^.
4. The resulting weights for each environment and mental health variable were recorded for each resample. The environment and mental health variables with non-zero loading above 90% across the resamples were selected and retained as stable variables in subsequent analyses.
5. We then re-applied the sCCA algorithm to the data, without sparsity constraints, on the stable urban living environment and mental health measures in the training dataset. The canonical correlation value between urban living environment and mental health measures were recorded.
6. We then permuted the training data, and repeated steps (2)-(5). This was done for 10,000 different permutations of the training data labelling. In each case, we recorded the canonical correlation value between urban living environment and mental health measures. In this way, we built up a permutation distribution to assess the significance of the relationship between urban living environment and mental health measures in the experimental labelling, within the training dataset.
7. We then applied the trained model to the test dataset to produce canonical correlates of urban living environment and mental health measures. We recorded associations for training and testing dataset.
8. We then randomly permuted the data rows in the test dataset and recalculated correlation values between urban living environment and mental health canonical correlates. We recorded associations between urban living environment and mental health correlates, for each of 10,000 permutations of the experimental labelling. False Discovery Rate (FDR) correction was used to control for multiple testings and a *P*_*FDR*_<0.05 was considered statistically significant.
9. After determining the significance of the first canonical correlate, we remove the effect of the first set of canonical vectors by projection deflation as we did before^19^, and repeat the sCCA analysis between the remaining urban living environment and mental health variables. These steps were iterated until the resulting canonical correlate were not significant any more.
10. To ensure reliability and reproducibility, we undertook further analyses including: a. sample composition using bootstrapping with replacement and random resampling; b. the effect of sex (Supplemental Methods).

Finally, we calculated the canonical correlations coefficient between the urban living environment canonical variables (which we refer to as ‘urban environmental profile’) and mental health canonical variables (referred to as ‘mental illness symptom group’), as well as structure coefficient value (*r*_*s*_) of individual variables

### GWAS analyses of the mental health symptoms groups that related to urban environment profile

For the significant mental illness symptom group that correlated with urban living environment profile from sCCA results, we conducted a genome-wide association analysis (GWAS) of the corresponding mental illness symptom group in 85,348 participants with complete genomic, urban environment and mental health data (Table 1). Using BGENIE v1.2 (https://jmarchini.org/bgenie/), we fit an additive model of association at each variant, using expected genotype count (dosage) from the imputed genetic data. The covariates included age, gender, assessment centre, processing batch and the top 10 ancestry principal components. Bonferroni *Pc*<0.05 (uncorrected *P*<0.05/139,187,27×numbers of significant mental illness symptom group from Scca results) was considered as a statistically significant threshold. We employed the FUMA online platform (https://fuma.ctglab.nl/) to fine mapping to genes and perform annotation of significant SNPs of each GWAS analysis. All SNPs with genome-wide significance were mapped to genes based on physical distance. Each SNP was mapped to the nearest gene in the human reference assembly (GRCh37/hg19).

### Gene-set enrichment analysis

To better understand the biological function of fine-mapping genes associated with mental health components, these genes were functionally annotated using ToppGene portal (https://toppgene.cchmc.org/) to identify significant enrichments for gene ontology (GO). The Benjamini and Hochberg method for false discovery rate (FDR-BH correction) (*q*_*c*_ < 0.05) was applied to correct for multiple comparisons. The default full reference gene list of each category in ToppGene were used as background gene set.

### Replication ananlysis

To replicate the multivariate relation between urban living environment and mental health, we applied the sCCA analysis in an independent dataset of 15,050 participants with complete environmental, mental health and neuroimaging data from UKB-NI dataset. Again, we used a training dataset (n=13,545, 90%) and a test dataset (n=1,505, 10%), a re-sampling method to ensure variable stability (with a threshold of 90% for non-zero weights from re-sampled data to consider as stable variables) and permutation tests to assess the significance of the results (10,000 times) as we used in the discovery sCCA analysis. Next, we independently replicated the significant SNPs associated with symptom groups of mental illness surviving from the discovery GWAS analysis (UKB-nonNI dataset) in an independent 8,726 participants of UKB-NI dataset at Bonferroni *Pc*<0.05 (uncorrected *P*<0.05/the numbers of all significant SNPs of GWAS from discovery analysis). Then we calculated the corresponding genes scores as the same way we did in the discovery analysis. And finally we independently validated the associations between genes scores and symptoms group of mental illness in the UKB-NI dataset.

### Brain volume changes underlying the associations between urban living environment and mental health

To investigate neurobiological mechanisms underlying the associations between urban living environment and mental health, we carried out a multiple sparse canonical correlation analysis (msCCA) between the environmental component, brain volume measures and the mental health component. This analysis was conducted in an independent sample of 15,050 participants with environmental, mental health and neuroimaging data from UK Biobank. Again, we used a training dataset (n=13,545, 90%) and a test dataset (n=1,505, 10%), a re-sampling method to ensure variable stability (with a threshold of 85% for non-zero weights from re-sampled data to consider as stable variables) and permutation tests to assess the significance of the results (10,000 times). An in-house Matlab script based on our previous work^19^ was employed for this analysis. Finally, the brain volume canonical variables (refered to as brain component), the canonical correlations coefficient between environment, brain and mental health canonical variables, structural loadings and weights of corresponding brain volume variables were calculated.

### Moderated mediation analysis between urban living enviroment, brain volume and mental health correlates modulated by genomics

The above analysis was carried out separately to identify the associations of mental health symptoms with urban living environment, genetic variation and brain volume, leaving the complex associations of urban living, genomics, brain and mental health unexplored. To formally test whether urban living environment-mental health relationship can be mediated by brain volume and modulated by genetics, we carried out a modulated mediation analysis in 8,726 participants. Moderated mediation analysis is an extension of mediation analysis, which is a valuable technique for assessing whether an indirect effect is conditional on a moderating variable. The basis of moderation and mediation effect were integrated into a combined model of moderated mediation within a linear regression framework. Finally, genomics was defined as modulated variables, the urban living environment correlate was defined as an independent variable, the brain volume correlates a mediator variable and the mental health correlates a dependent variable.

In moderated mediation analyses, all indirect effects are estimated in one multiple regression analysis with independent variable and all mediators as predictor variables. This means that the indirect effect of one mediator was estimated when the other mediators are taken into account. We used bootstrapping to assess the significance of the mediation effect. After 5,000 bias-corrected bootstrapping, we estimated the distribution of the indirect effect and calculate its 95% confidence intervals (CI). If zero does not fall between the resulting 95% confidence interval of the bootstrapping method, we confirmed the existence of a significant mediation effect (*P*<0.05). It should be emphasized that in the multiple mediation analysis of this study, mediators and dependent variables were measured contemporaneously, thus not allowing establishment of any causal directionality. Explained mediation effect (EME) and 95% CI were reported for moderated mediation analyses. Confounding factors were controlled in the moderated mediation model. The moderated mediation analysis with a nonparametric bootstrap method was conducted using the *mediation* R package^77^.

## Data Availability

All data produced in the present study are available upon reasonable request to the authors

