## Supplementary Materials for "Environmental profiles of urban living relate to regional brain volumes and symptom groups of mental illness through distinct genetic pathways"

### Supplementary Methods

#### Overview of analyses

In 141,325 participants without neuroimaging data (UKB-nonNI), we identified environmental profiles correlated to symptom groups of emotional problems by splitting our data in training and test datasets, and applying bootstrapping with replacement and random resampling. Next, we carried out genome-wide association analysis (GWAS) analyses of the symptom groups identified in 85,348 participants with complete genomic, environment and mental health data from the UKB-nonNI dataset. The dataset with neuroimaging data (n=15,050, UKB-NI) was used for independent replication of the multivariate relation between urban living environment, genes and mental health, as well as for additional neuroimaging analyses. We analysed relations between the environmental profiles, regional brain volume and the emotional symptom groups applying msCCA. Again, we used a split design with a training and a test dataset. We then investigated the mediation of the effect of the environmental profiles on emotional symptom groups by regional brain volume and its genetic moderation in 8,726 participants with complete genomic, environment, brain volume and mental health data in UKB-NI.

#### Street network morphometric

Physical accessibility of street network was modelled through spatial Design Network Analysis (sDNA). sDNA is a sophisticated technique of urban network analysis that has evolved from conventional network analyses and employs street network links as the fundamental unit of computation[1]. The Ordnance Survey

Mastermap Integrated Transport Network (OSM ITN) was subjected to automated cleaning in sDNA Prepare Tooland subsequent modelling produced a suite of 18 different indices of street-network accessibility (Supplementary Table 2). These measure link, centrality, radial, detour, shape characteristics of urban morphology captured at micro (neighbourhood)-, meso (city)- and macro (regional)-level encompassing 19 different catchment radius (400-50,000 m). Here we only used the measures with catchment radius of 400m (Supplementary Table 2). These are generated in sDNA for all links in the urban road network covering the entire UKB cohort members and the metrics for a street links containing a UKB respondent's dwelling were added to the respondent's BE profile. The detailed description of these street network morphometric measures are shown in Supplementary Table 3.

**Replacement and random resampling**

In addition to random permutations and evaluating the loadings of each feature on an independent test sample, we still undertook multiple steps to evaluate the robustness and reliability of the results in sCCA analysis between urban living environment profiles and mental illness symptoms.

(1) sCCA stability: We assessed the stability of each sCCA analysis by rerunning the algorithm in 1000 randomly generated subsets, each containing 10% to 150% of the training dataset in 10% increments and recalculating the canonical correlation between urban living environment profiles and mental health symptoms (Supplementary Table 6).

(2) Random resampling: We resampled 90% of the training dataset 1000 times, reran the sCCA algorithm and calculated the canonical correlation between the resulting feature loadings in the remaining 10% of the training dataset each time.

(3) Effect of sex: To evaluate whether there was similarity of the original sCCA modes between males and females, we calculated the canonical correlation between urban living environment and mental health symptoms in males and females,

respectively. The canonical correlation of males and females were then calculated for all 3 significant modes in both training and test datasets (Supplementary Table 7).

#### Supplementary Results

##### Gene-set enrichment analysis highlight relevant neurobiology

We performed bioinformatic analyses to explore mechanism underlying the genes associated with the three mental illness symptom groups using TopGene[2] (Figure 3e and Supplementary Tables 12). The Benjamini and Hochberg method for false discovery rate (FDR-BH correction) ($qc<0.05$) was applied to correct for multiple comparisons. The reference gene list was used as default setting. In the 43 genes associated with the affective symptom group, the results suggested enrichment in genes expressed in brain and pituitary tissues and involved in nervous system development, in particular neuron differentiation ($qc = 5.63 \times 10^{-3}$) and regulation of axonogenesis ($qc = 5.64 \times 10^{-3}$) (Fig. 3c and Supplementary Table 12), namely in the cerebral cortex during prenatal development (Supplementary Table 12). We also found over-representation in the molecular function of coticotropin-releasing hormone (CRH) receptor activity ($qc = 4.37 \times 10^{-4}$) and CRH binding ($qc = 8.71 \times 10^{-4}$), as well as coticotropin-releasing factor receptor activity ($qc = 4.37 \times 10^{-4}$) (Supplementary Table 12). These functional annotations suggest that the CRH signalling is essential for shaping environment-related affective symptoms presumably by regulating stress and immune response[3,4].

In the 11 genes related to the anxiety symptom group, we found enrichment in molecular function of small nucleolar ribonucleoprotein (snoRNP) complex binding, which may influence anxiety disorders directly or through serotonin receptors (Supplementary Table 12). In the 6 genes related to the emotional instability symptom group, we found significant enrichment in the cerebellar granular layer development process ($qc = 6.42 \times 10^{-4}$) (Supplementary Table 12). The detailed gene-set enrichment items of affective, anxiety and emotional instability symptom groups are shown in Supplementary Table 12.

#### Gene score calculation

We created a score for the genes associated with each mental illness symptom group, using plink 2.0[5] with default parameter settings adjusted for linkage disequilibrium. Specifically, the clump-p1 indicated GWAS *P*-value threshold for a SNP to be included as an index SNP, which is set to 1 such that all SNPs are include for clumping. The clump-r2 is set to 0.5, indicating SNPs having r2 higher than 0.5 with the index SNPs will be removed. The clump-kb is set to 250kb, indicating SNPs within 250k of the index SNP are considered for clumping. The score of each gene is then calculated as the sum of the count of risk alleles multiplied by the corresponding beta value from GWAS across the remaining index SNPs. Thus, we generated 43 genes scores associated with the affective symptom group, 11 genes scores for the anxiety symptom group and 6 genes scores for the emotional instability symptom group.

#### Supplementary Tables

**Supplementary Table 1. Urban living environmental variables and categories.**

| Area | Category | No. of subcategory | No. of items | Sample size (n) |
|---|---|---|---|---|
| Traffic | Traffic | 1 | 7 | 502,617 |
| Residential pollution | Air pollution | 1 | 6 | 502,617 |
| | Sound pollution | 1 | 5 | 502,617 |
| Greenspace and coastal proximity | Natural environment | 1 | 1 | 497,519 |
| | Greenspace | 1 | 1 | 440,851 |
| | Domestic garden | 1 | 1 | 440,851 |
| | Domestic water | 1 | 1 | 440,851 |
| | Distance from home to coast | 1 | 1 | 497,519 |
| IMD | EIMD* | 1 | 1 | 424,419 |
| | SIMD* | 1 | 1 | 424,419 |
| | WIMD* | 1 | 1 | 424,419 |
| Urban morphometry | Building Class | 1 | 3 | 317,095 |
| | Terrain | 2 | 2 | 430,832 |
| | Destination Accessibility | 9 | 33 | 423,998 |

| | | | | |
|---|---|---|---|---|
| | Greenness | 2 | 2 | 430,832 |
| | Landuse Density | 25 | 46 | 424,028 |
| | Street Network Accessibility | 5 | 18 | 423,673 |
| Total | 15 | 53 | 128 | 216,341 |

*EIMD, England Indices of multiple deprivation; SIMD, Scotland Indices of multiple deprivation; WIMD, Wales Indices of multiple deprivation.*

*Note: each participant only has one IMD score based on their residential locations in England, Scotland and Wales.*

**Supplementary Table 2. Detailed urban living environmental variables. (see Excel)**

**Supplementary Table 3. Description of sDNA modelled street accessibility indices**

| Variable name | Description |
|---|---|
| **Link characteristics:These measures describe the characteristics of individual links in the network** | |
| 1. Link Connectivity | The number of link ends that an individual link is connected to at its end points |
| 2. Link Length | Length of the individual link in the network |
| 3. Link Angular Curvature | The cumulative angular change while traversing the full length of a link in degrees |
| **Centrality analysis:These set of measures owe their origin to the graph theory. The associations between urban morphology and the social phenomena dependent on it are essentially captured by indices of relationality in the graphs. The notion of accessibility captured by these measures acts to formally elucidate how network morphology influences individual activity behaviours and drives various socio- economic processes. They indicate the centrality of a vertex within a graph** | |
| 1. Mean Angular Distance | In graphical terminology, also called as the closeness centrality/accessibility. It is an indicator of the degree of difficulty, on average, of navigating to all possible destinations within a specified radius from each given link. This is weighted by the link length |
| 2. Network Quantity Penalized for Distance | This is an improved measure of the conventional closeness centrality and takes in to account the effects of network quantity. For each link within a specified radius, it takes the network quantity (defined link length) and divides it by the difficulty of access (angular). This is weighted by the link length. |
| 3. Betweenness | In graphical terminology, also called as the betweenness centrality or path overlap or through-movement potential. It is indicative of how often a given link is used for a journey within a defined radius. Measured as the sum of geodesics that pass through a link for a journey within a defined radius. This has been weighted by origin-destination link length. |

| | | |
|---|---|---|
| 4. | Two Phase Betweenness | This is betweenness weighted by a two-step floating catchment model. Measured as the sum of geodesics that pass through a link for a journey within a defined radius weighted by the proportion of network quantity accessible from geodesic origin that is represented by geodesic destination |
| 5. | Two Phase Destination assignment | This is the total flow to each destination under the two phase betweenness model. In other words, it is similar to the two phase betweenness, but measured for the destination of each geodesic only |

**Simple radial measures:These measures pertain to the characteristics of the links within a specified network radius.**

| | | |
|---|---|---|
| 1. | Links | The number of network links within a specified network radius |
| 2. | Length | The total network length within a specified network radius |
| 3. | Angular Distance | Sum of angular distance of each individual link within a specified radius |
| 4. | Weight | Total weight within a specified radius. Weights have been specified with respect unit of network length (in length weighted analysis) |
| 5. | Mean Geometric Length | Mean of the angular geodesic Euclidean length within a specified radius. This has been weighted by the origin to destination link length |

**Network detour analysis: Measure the network severance by comparing the hypothetical crow fly distance to actual network distance. It is an indicator of the extent of deviation of the network from the most direct path.**

| | | |
|---|---|---|
| 1. | Mean Crow Flight Distance | Mean of the crow flight distance between a link and all the links within a defined radius. This is weighted by the link length |
| 2. | Diversion Ratio | Mean of the ratio of actual geodesic length to the crow flight distance for all geodesics within a defined radius. This is weighted by the link length. Indicative of the degree of deviation of the actual paths from the crow flight path |

**Network shape: Measure of network efficiency in terms of the spatial footprint of the street network in urban space.**

| | | |
|---|---|---|
| 1. | Convex Hull Area | Area of the convex hull containing all the origins and destinations within a defined radius. It is an indicator of the network footprint or the spatial spread of the street network in the urban space. |
| 2. | Convex Hull Perimeter | Length of perimeter of the convex hull containing all the origins and destinations within a defined radius. |
| | | 400, |
| 3. | Convex Hull Maximum Radius | Maximum radius of the convex hull measured as the crow flight distance from the centre of the origin link to the furthest point on the convex hull of a defined radius |
| 4. | Convex Hull Bearing | Compass bearing of the line of maximum radius of convex hull of a defined radius, measured in degrees. It indicates the direction in which one can travel furthest from the origin link, while staying inside the network radius. |
| 5. | Convex Hull Shape Index | Measures the degree of uniformity of the network in all directions. It is measured as the square of the hull perimeter divided by 4π times the hull area. Ranges from 1 in case of a circle to higher values, with higher indicating non-uniformity across all directions. |

**Supplementary Table 4. Details of 21 symptoms of mental illness in UK biobank**

| Field ID | Variable name | Question | Sample size (n) |
|---|---|---|---|
| 1920 | Mood swings | Does your mood often go up and down? | 488,298 |
| 1930 | Miserableness | Do you ever feel 'just miserable' for no reason? | 492,219 |
| 1940 | Irritability | Are you an irritable person? | 478,082 |
| 1950 | Sensitivity hurt feelings | Are your feelings easily hurt? | 486,281 |
| 1960 | Fed up feelings | Do you often feel 'fed-up'? | 490,044 |
| 1970 | Nervous feelings | Would you call yourself a nervous person? | 487,683 |
| 1980 | Worrier /anxious feelings | Are you a worrier? | 487,727 |
| 1990 | Tense /highly strung | Would you call yourself tense or 'highly strung'? | 482,778 |
| 2000 | Worry too long | Do you worry too long after an embarrassing experience? | 480,269 |
| 2010 | Suffering from nerves | Do you suffer from 'nerves'? | 481,980 |
| 2020 | Loneliness and isolation | Do you often feel lonely? | 492,739 |
| 2030 | Guilty feelings | Are you often troubled by feelings of guilt? | 487,212 |
| 2040 | Risk taking | Would you describe yourself as someone who takes risks? | 482,170 |
| 2050 | Frequency of depressed mood | Over the past two weeks, how often have you felt down, depressed or hopeless? | 478,435 |
| 2060 | Frequency of unenthusiasm | Over the past two weeks, how often have you had little interest or pleasure in doing things? | 482,800 |

| | | | |
|---|---|---|---|
| 2070 | Frequency of tenseness | Over the past two weeks, how often have you felt tense, fidgety or restless? | 480,413 |
| 2080 | Frequency of tiredness | Over the past two weeks, how often have you felt tired or had little energy? | 485,357 |
| 2090 | Seen a doctor (GP) | Have you ever seen a general practitioner (GP) for nerves, anxiety, tension or depression? | 501,704 |
| 2100 | Seen a psychiatrist | Have you ever seen a psychiatrist for nerves, anxiety, tension or depression? | 501,704 |
| 6145 | Illness, injury, bereavement and stress | In the last 2 years have you experienced any of the following? | 497,926 |
| 20217 | Neuroticism score | Based on 12 neurotic behaviour domains | 401,652 |

**Supplementary Table 5. Structure coefficient of urban living environment profile and symptoms of mental illness in sCCA-regression analysis (see Excel)**

**Supplementary Table 6. Multiple comparisons of canonical correlation in each resampling of training dataset.**

| Category | Mean | Mean difference | 95% CI | 95% CI | *P* value |
|---|---|---|---|---|---|
| 100% vs. 10% | 0.2156 | -0.009357 | -0.01088 | -0.007837 | ***<0.0001*** |
| 100% vs. 20% | 0.2120 | -0.005734 | -0.007255 | -0.004214 | ***<0.0001*** |
| 100% vs. 30% | 0.2094 | -0.003153 | -0.004673 | -0.001632 | ***<0.0001*** |
| 100% vs. 40% | 0.2082 | -0.001956 | -0.003477 | -0.0004359 | ***0.0037*** |
| 100% vs. 50% | 0.2074 | -0.001175 | -0.002695 | 0.0003456 | 0.2320 |
| 100% vs. 60% | 0.2071 | -0.0008983 | -0.002419 | 0.0006221 | 0.5591 |
| 100% vs. 70% | 0.2072 | -0.0009570 | -0.002477 | 0.0005635 | 0.4768 |
| 100% vs. 80% | 0.2070 | -0.0007446 | -0.002265 | 0.0007758 | 0.7790 |
| 100% vs. 90% | 0.2065 | -0.0002455 | -0.001766 | 0.001275 | 0.9994 |
| 100% vs. 110% | 0.2061 | 0.0001150 | -0.001406 | 0.001635 | 0.9997 |
| 100% vs. 120% | 0.2065 | -0.0002426 | -0.001763 | 0.001278 | 0.9994 |
| 100% vs. 130% | 0.2060 | 0.0002525 | -0.001268 | 0.001773 | 0.9993 |
| 100% vs. 140% | 0.2062 | 2.210e-006 | -0.001518 | 0.001523 | >0.9999 |
| 100% vs. 150% | 0.2062 | -2.253e-005 | -0.001543 | 0.001498 | >0.9999 |

**Supplementary Table 7. Canonical correlation between urban living environmental profile and symptom group of mental illness in male and females.**

| | Training dataset | | Test dataset | | |
|---|---|---|---|---|---|
| | ***cc*** **value** | $P_{permutation}$ **value** | ***cc*** **value** | $P_{permutation}$ **value** | **FDR** ***P*** **value** |
| **Female** | | | | | |
| Affective symptom group | 0.205 | ***P<0.001*** | 0.185 | ***P<0.001*** | ***P<0.001*** |
| Anxiety symptom group | 0.120 | ***P<0.001*** | 0.079 | ***P<0.001*** | ***P<0.001*** |
| Emotional instability symptom group | 0.059 | ***P<0.001*** | 0.010 | 0.179 | 0.277 |
| **Male** | | | | | |
| Affective symptom group | 0.208 | ***P<0.001*** | 0.182 | ***P<0.001*** | ***P<0.001*** |
| Anxiety symptom group | 0.106 | ***P<0.001*** | 0.109 | ***P<0.001*** | ***P<0.001*** |
| Emotional instability symptom group | 0.060 | ***P<0.001*** | 0.045 | ***P<0.001*** | ***0.002*** |

**Supplementary Table 8. Replication of multivariate relation between urban living environment and mental illness symptoms group in the independent UKB-NI dataset (see Excel)**

**Supplementary Table 9. 2,983 significant GWAS associations between SNPs and affective symptoms group at Bonferroni *Pc*<0.05 (see Excel)**

**Supplementary Table 10. 29 significant GWAS associations between SNPs and anxiety symptoms group at Bonferroni *Pc*<0.05 (see Excel)**

**Supplementary Table 11. 6 significant GWAS associations between SNPs and emotional instability symptoms group at Bonferroni *Pc*<0.05 (see Excel)**

**Supplementary Table 12. Gene-set enrichment analysis results in gene ontology of affective, anxiety and emotional instability symptoms group (see Excel)**

**Supplementary Table 13-15. Replications of GWAS analysis of mental illness symptom groups in UKB-NI dataset (see Excel)**

**Supplementary Table 16. Replications of genes score associated with mental illness symptom groups (see Excel)**

**Supplementary Table 17. msCCA loadings for brain volumes simultaneously associated with urban living environment profiles and mental illness symptoms groups**

| Mental health | Brain area | Loading |
|---|---|---|
| Affective symptom group | Left Superior Frontal Gyrus | -0.457 |
| | Right Superior Frontal Gyrus | -0.429 |
| | Right Cerebellum VIIIa | -0.438 |
| | Right Cerebellum VIIb | -0.332 |
| | Right Cerebellum VIIIb | -0.328 |
| | Right Crus I Cerebellum | -0.319 |
| | Left Cerebellum VIIIa | -0.296 |
| | Right Crus II Cerebellum | -0.287 |
| | Left Amygdala | -0.278 |
| | Left Cerebellum VIIb | -0.256 |
| | Right Accumbens | -0.246 |
| | Right Occipital Fusiform Gyrus | -0.242 |
| | Right Frontal Pole | -0.239 |
| Anxiety symptom group | Right VIIIa cerebellum | -0.396 |
| | Right crus I cerebellum | -0.351 |
| | Right V cerebellum | -0.301 |
| | Left VIIIb cerebellum | -0.300 |
| | Right VIIIb cerebellum | -0.291 |
| | Left crus I of cerebellum | -0.289 |
| | Left Juxtapositional lobule cortex | -0.287 |
| | Left VI cerebellum | -0.278 |
| | Right amygdala | -0.277 |
| | Left VIIIa cerebellum | -0.274 |
| | Left inferior frontal gyrus | -0.250 |
| Emotional instability symptom group | Left Precentral Gyrus | -0.391 |
| | Right Frontal Pole | -0.344 |

| | | |
|---|---|---|
| | Left Frontal Pole | -0.339 |
| | Right Precentral Gyrus | -0.321 |
| | Right Amygdala | -0.292 |
| | Left Amygdala | -0.278 |
| | Left Insular Cortex | -0.276 |
| | Left Lateral Occipital Cortex | -0.275 |
| | Left Central Opercular Cortex | -0.270 |
| | Right Frontal Orbital Cortex | -0.270 |
| | Left Crus I Cerebellum | -0.269 |
| | Left Postcentral Gyrus | -0.268 |
| | Right Insular Cortex | -0.262 |

**Supplementary Figures**

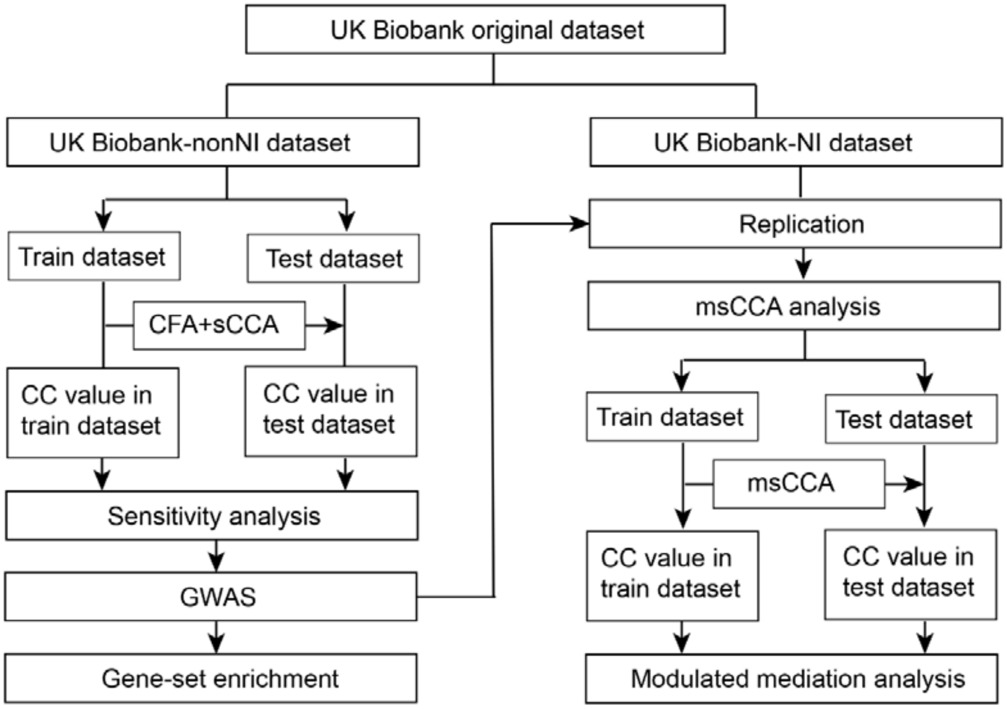

***Supplementary Figure 1. A schematic summary of the study design.*** *UK biobank-non NI dataset and NI dataset, participants from UK Biobank with complete urban living environmental data and mental health data (n=156,375) were divided into datasets without neuroimaging data (UK biobank-non NI dataset, n= 141,325) and with neuroimaging data (UK biobank-NI dataset, n=15,050). CC value, canonical correlation value; CFA, confirmatory factor analysis; GWAS, genome-wide association analysis; msCCA, multiple sparse canonical correlation analysis; sCCA, sparse canonical correlation analysis.*

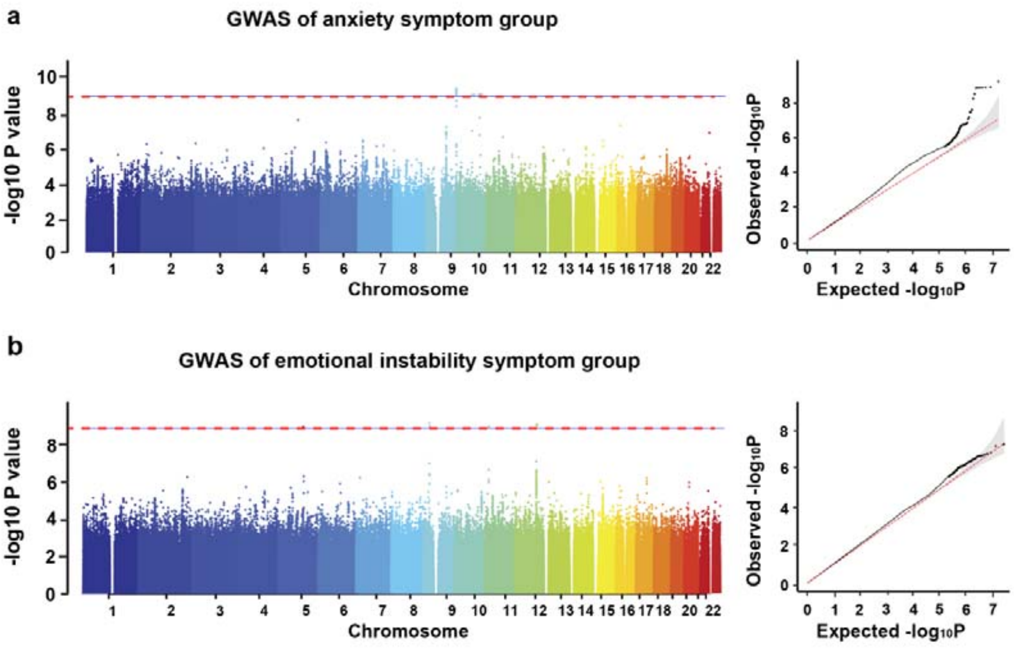

***Supplementary Figure 2. GWAS analysis of the canonical variates of the anxiety (a) and emotional instability (b) symptom groups in 85,348 participants with complete genetic, urban environment and mental health data in UKB-nonNI datasets.*** *a. Left. Manhattan plot of GWAS analysis of canonical variates of the anxiety symptom groups. We found significant associations of the anxiety symptom group with 29 SNPs at Bonferroni Pc<0.05; Right. Quantile-quantile plot of GWAS analysis. b. Manhattan plot of GWAS analysis of canonical variates of the emotional instability symptom groups. There were 6 significant genetic associations at Bonferroni Pc<0.05; Right. Quantile-quantile plot of GWAS analysis.*
